# Epidemiology of Syphilis infections among pregnant women in Tanzania: Analysis of the 2020 national representative sentinel surveillance

**DOI:** 10.1101/2023.04.16.23288656

**Authors:** Bruno Sunguya, Erick Mboya, Mucho Mizinduko, Belinda Balandya, Amon Sabasaba, Davis Elias Amani, Doreen Kamori, George Ruhago, Rebecca Mkumbwa, Prosper Faustine, Werner Maokola, Veryeh Sambu, Jeremiah Mushi, Mukome Nyamuhagata, Boniphace S. Jullu, Amiri Juya, Joan Rugemalila, George Mgomella, Sarah Asiimwe, Andrea B. Pembe

**Affiliations:** Muhimbili University of Health and Allied Sciences, Dar es salaam, Tanzania; Ministry of Health, Dodoma, Tanzania; St. Francis University College of Health and Allied Sciences, Morogoro, Tanzania; Tanzania Field Epidemiology and Laboratory Training Program, Dar es salaam, Tanzania; Muhimbili National Hospital, Dar es Salaam, Tanzania; Centre for Diseases Control, Tanzania; Global Fund, Tanzania Country Team

**Author notes:** Deceased (Boniphace S. Jullu).

**Keywords:** Syphilis, Pregnancy, Antenatal care, Tanzania

## Abstract

**Background:** Syphilis has detrimental effects on the health of the mother and that of the child when pregnant. Understanding its local epidemiology is essential for policies, planning, and implementation of targeted preventive interventions. Using data from the 2020 National Sentinel Surveillance of pregnant women attending antenatal clinics (ANCs) in Tanzania we determined the prevalence and determinants of Syphilis among pregnant women in Tanzania mainland.

**Methodology:** The ANC surveillance was conducted in 159 ANC sites from all 26 regions of Tanzania’s mainland from September to December 2020. It included all pregnant women older than 14 years on their first ANC visit in the current pregnancy during the survey period. Counselling for Syphilis was done using standard guidelines at the ANC and testing was done using rapid SD Bioline HIV/Syphilis Duo test kits. Analysis was done using both descriptive statistics to determine the prevalence and characteristics of syphilis, whereas, logistic regressions were used to examine the independent association between syphilis and dependent variables.

**Results:** A total of 38,783 women [median age (Interquartile range (IQR)) =25 (21-30) years] participated in the surveillance. Of them, 582 (1.4%) tested positive for Syphilis. A wide regional variation was observed with the highest burden in Kagera (4.5%) to the lowest burden in Kigoma (0.3%). The odds of Syphilis infections were higher among older women and those with no formal education. Compared with primigravids, women with 1-2, those with 3-4 and those with more than four previous pregnancies had 1.8 (aOR=1.8, 95% CI: 1.2-2.5), 2.1 (aOR=2.1, 95% CI: 1.4-3.1) and 2.6 (aOR=2.6, 95% CI: 1.7-3.9) higher odds of syphilis infection respectively.

**Conclusion:** Syphilis is still prevalent among pregnant women in Tanzania with a wide regional disparity. Efforts to prevent new infections, screen pregnant women, and treat those infected should be strategized to include all regions and renewed emphasis in regions with high burden, and importantly among women who are multipara, with the low level of education, and advanced age.

## Introduction

Caused by *Treponema pallidum*, Syphilis has remained a detrimental sexually transmitted disease, more prevalent in low-and middle-income countries (LMICs), and can be transmitted from mother to child (Cohn *et al*., 2021; WHO, 2021). About 900,000 pregnant women were infected with Syphilis and over 660,000 cases of congenital syphilis in 2016 were noted in 2016 alone (Korenromp *et al*., 2019). The WHO African region has the highest prevalence of Syphilis (1.5%) among pregnant women compared to other regions in the world (Cohn *et al*., 2021). Within the continent, the East African region has been reported to be the region with the highest prevalence of syphilis in pregnancy (Hussen and Tadesse, 2019). Efforts to address the burden of syphilis has resulted into a steady decline of its prevalence in Tanzania, especially among pregnant women from 8.3% in 2000 to 2% in 2011 (Swai *et al*., 2006; Manyahi *et al*., 2015, 2017).

Syphilis has detrimental effects on the health of the mother and that of the child when pregnant. In 2019, over half of Syphilis cases in pregnancy suffered adverse birth outcomes including early fetal deaths, stillbirths, neonatal deaths, pre-term birth or low birth weights, and clinical congenital syphilis (Korenromp *et al*., 2019). Additionally, HIV and Syphilis infections interact and co-infection during pregnancy increases the risk of vertical transmission of HIV coupled with adverse outcomes of pregnancy (Mwapasa *et al*., 2006; Shava, Moyo and Zash, 2019).

Owing to the unprecedented effect of the disease, especially during pregnancy and therefore birth outcomes, prevention efforts are of utmost importance. These efforts should be coupled with those of prevention of Mother to Child Transmission of HIV and that of other STIs. The effects of such efforts need to be verified through quality data to inform the national response, determining areas and regions with remaining challenge, populations more at risk, and characteristics of the burden in the country. The last of such survey is one decade old. There is therefore a gap for comprehensive and updated information about Syphilis infections in pregnancy at the national and subnational levels in Tanzania thus rendering difficulty ascertaining the burden accurately.

For programs and stakeholders to efficiently plan, and implement interventions to reduce morbidity and mortality from these infections, they need accurate information about the distribution of the infections at the national and sub-national levels. Such interventions include prevention of new cases, ensuring availability and accessibility of testing services, initiation and retention to treatment, safe delivery and post-natal prophylaxis as well as early diagnosis of congenital syphilis (Cock *et al*., 2015; Hurst, 2015; Drake *et al*., 2019; WHO, 2021). Recognizing the need for updated and comprehensive information on syphilis infections among pregnant women in Tanzania, and using the data from the 2020 antenatal HIV and Syphilis sentinel surveillance in Tanzania mainland, we determined the prevalence of syphilis and the factors associated with the infection among pregnant women in Tanzania mainland.

## Methodology

### Surveillance duration and site selection

Data for the Antenatal Clinic (ANC) HIV & Syphilis Sentinel Surveillance (HSS) was collected from September to December 2020 in all 26 regions of Tanzania mainland. A total of 159 antenatal clinics (ANCs) providing Prevention of Mother to Child Transmission (PMTCT) services were included in this surveillance. In Tanzania, ANC coverage of at least one visit is more than 98%, and Syphilis testing services are integrated into ANCs (WHO, 2022). All pregnant women are counselled, and routinely offered Syphilis test using a national guideline and as a standard of care. Those who test positive are immediately initiated on treatment in line with the national guidelines (MoH, 2013). In addition, women who opt out at their first ANC visit are informed that they can access Syphilis testing at any future visit if they change their minds (MoH, 2013). As for the previous HSS rounds, two urban, two semi-urban, and two rural sites were selected from each region, except for the Dar es Salaam region, where all six sites are classified as urban.

### Survey population

All pregnant women aged 15 years and above attending ANC on their first visit in the current pregnancy during the survey period were eligible for inclusion in the ANC HSS. However, only those who provided written consent and were willing to give a blood sample for routine Prevention of Mother To Child Transmission (PMTCT) of HIV and Syphilis testing were included in the surveillance. All sites enrolled ANC attendees who were making their debut visits. To achieve the targeted sample size, all sites carried out data collection continuously for three consecutive months.

### Training of research personnel

A group of national-level facilitators was trained on implementing the ANC HSS by the principal investigator and the co-investigators. These national-level facilitators then conducted zonal training workshops organized in five zones before the commencement of the survey. Participants included nurses attending ANC clients from all 159 sites and regional and district personnel involved in the survey implementation. The training content included a review of operational procedures and field protocols for ANC HSS based on routine data, including training on the updated ANC HSS data collection form and HIV and Syphilis testing practical. In addition, a team of national-level study staff was trained to monitor the quality of routinely collected PMTCT data and the quality of PMTCT HIV and Syphilis testing.

### Data collection

This surveillance was integrated into routine ANC activities where all women attending the ANC on a particular day were informed of the ongoing surveillance. The provider explained the study and its procedures, and later obtained written informed consent from the participant for each component of the survey (the interview, and routine rapid testing for HIV and Syphilis). In addition, a non-identifying unique survey ID code (HSS number) pre-printed on barcode stickers was assigned to each prospective survey participant found to be eligible.

The questionnaire used, was developed in English, translated into Swahili, and then translated back into English by a third party to verify the accuracy of the translation. It was administered by a trained RCH provider using an electronic tablet loaded with open data kit (ODK), a data collection software. It captured the HSS number, ANC number, age, marital status, gravidity, education level, residence, employment, and Syphilis results. Electronic data were uploaded to a password-protected server at the end of each day.

### Blood testing procedures

Following the interview, blood sample was drawn from all consented participants. Syphilis testing was conducted according to the national guidelines and the national rapid testing algorithm (NACP, 2019). Since the HSS was conducted for both HIV and Syphilis, the provider conducted HIV and Syphilis testing using rapid SD Bioline HIV/ Syphilis Duo test kits. The Ministry of Health (MOH) adopted the kit owing to its cost-effectiveness in testing for both HIV and Syphilis and enables broader Syphilis testing coverage (Lyamuya *et al*., 2009; Olugbenga *et al*., 2018). Each participant received their result in a post-test counselling session participants with positive Syphilis test results were treated according to National STI Management Guidelines. Syphilis test result data were recorded on a register and entered into an ODK form.

### Data management and analysis

Sociodemographic data and routine blood testing data collected were uploaded daily to a password-protected cloud storage system by each survey team member to allow for real-time data monitoring. To minimize data entry errors, questions in the ODK had prompts and checks for data validation and correction of identified errors.

Analysis was conducted using Stata Statistical software ver. 17 (College Station, TX: Stata Corp LLC.). The prevalence of pregnant women infected with Syphilis was calculated at the national and regional levels, disaggregated by demographic characteristics. The prevalence and the logistic regression estimates explained below considered the survey design of this surveillance (clustering at the health facility level and stratification by locality that is rural, semi-urban, and urban). The estimates were also weighed using inverse probability weights based on the total number of women who attended ANC during the study period at each region. Logistic regressions were used to examine the independent association between dependent variables, Syphilis infection, with independent variables such as age, marital status, parity, education level, source of income, and locality. Known confounders such as age and other factors with associations reaching p-value <0.2 at the univariate analysis were included in the multivariate model. The level of statistical significance was set at p<0.05.

### Ethical consideration

Study staff sought informed consent from potential participants before they engaged them in the survey. Pregnant women who did not consent to participate in the survey received routine ANC care as supposed. Participants were assured that their responses would be kept confidential and that no harm would come to them because of agreeing or not agreeing to participate in the survey. Approval to conduct this study was provided by the MUHAS Institutional Review Board (MUHAS-REC-07-2020-298). Strict protection ensured data security and confidentiality. The ANC HSS did not collect any personally identifiable information other than what was essential to meet the assessment objectives.

## Results

### Demographic characteristics

The 2020 survey included 39,516 pregnant women. Of them, 38,783 (98.1%), consented to participate in the surveillance and tested for Syphilis. Demographic characteristics were as follows: Their median age (inter quartile range (IQR)) was 25 (21–30) years, and 45.8% of them were between 15–24 years. About 78.5% were married, 60.9% had attained primary school education, 48.9% were housewives, and 44.6% had at most two previous pregnancies. One in four (26.3%) was a primigravida. Almost half of the participants, 48.6% were recruited from facilities located in urban areas. **Table 1**.

**Table 1:**
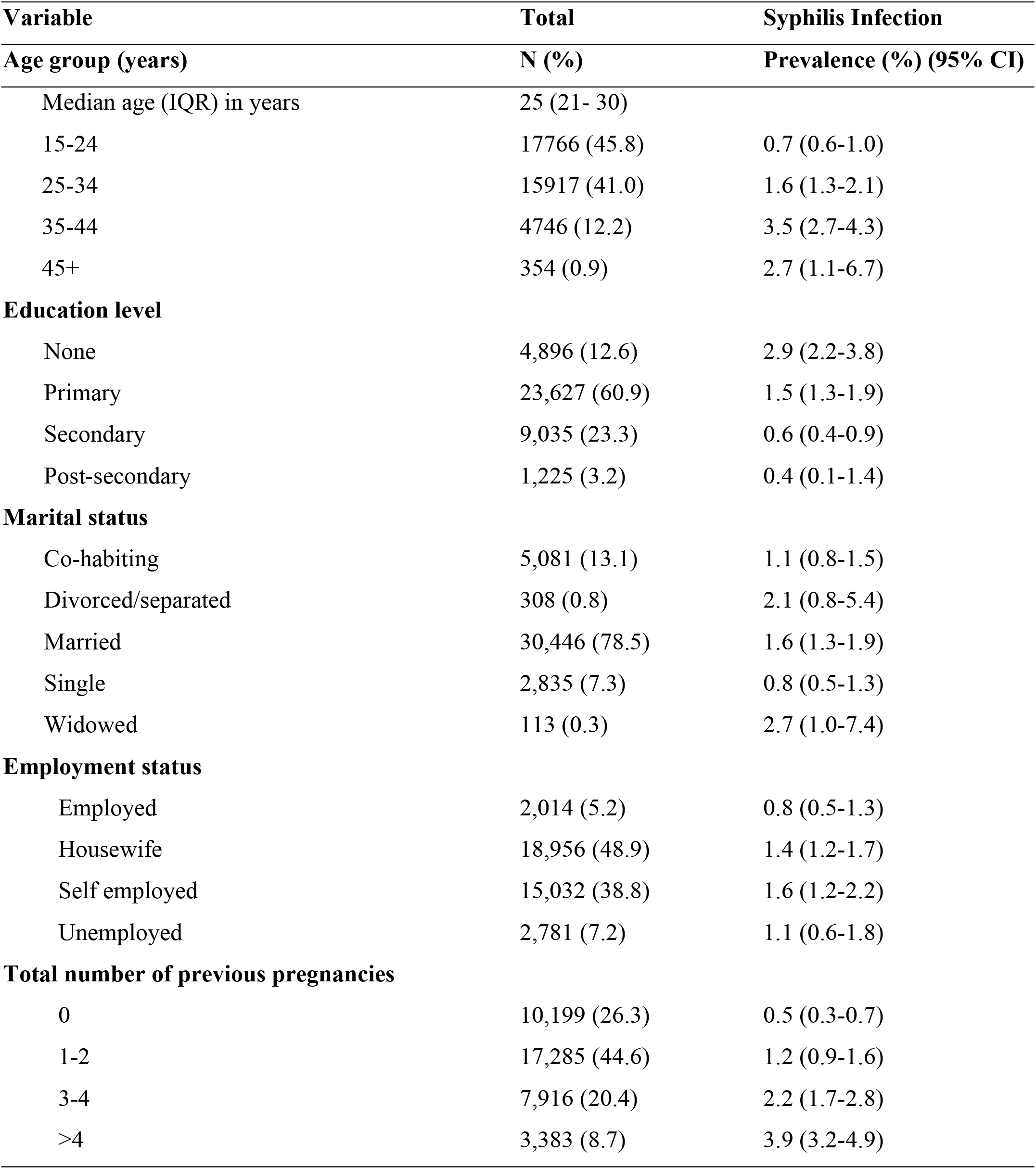
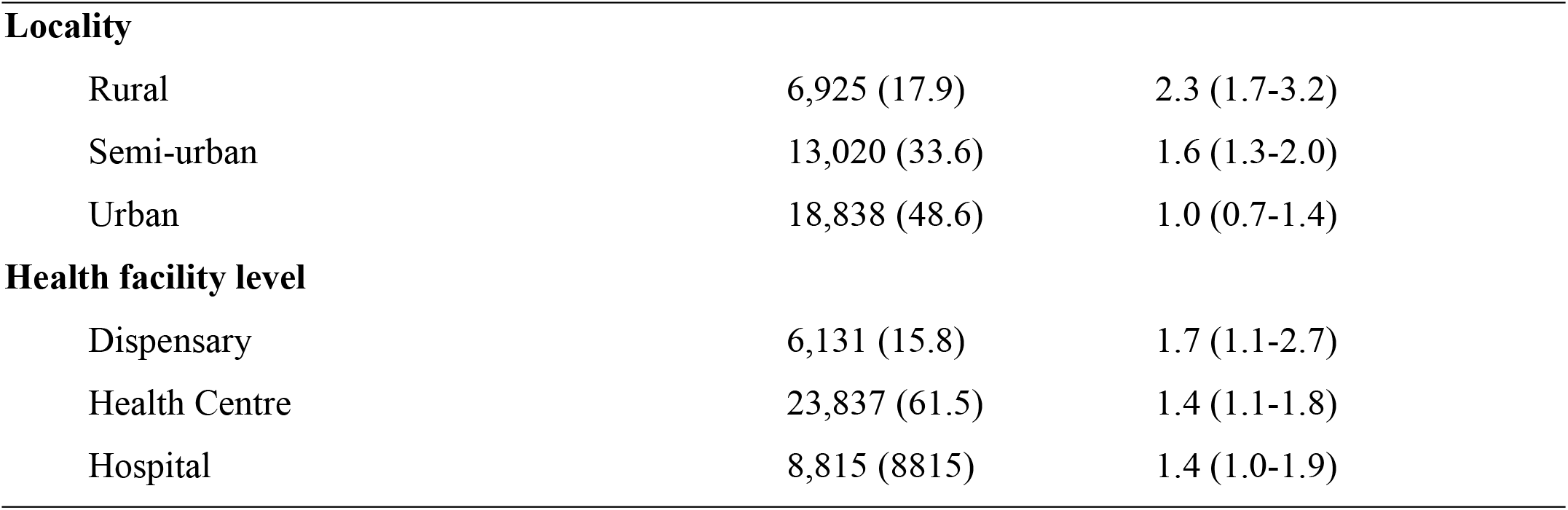
Demographic characteristics of participants and prevalence of Syphilis among pregnant women in Tanzania.

| **Variable** | **Total** | **Syphilis Infection** |
| --- | --- | --- |
| **Age group (years)** | **N (%)** | **Prevalence (%) (95% CI)** |
| Median age (IQR) in years | 25 (21- 30) |  |
| 15-24 | 17766 (45.8) | 0.7 (0.6-1.0) |
| 25-34 | 15917 (41.0) | 1.6 (1.3-2.1) |
| 35-44 | 4746 (12.2) | 3.5 (2.7-4.3) |
| 45+ | 354 (0.9) | 2.7 (1.1-6.7) |
| **Education level** |  |  |
| None | 4,896 (12.6) | 2.9 (2.2-3.8) |
| Primary | 23,627 (60.9) | 1.5 (1.3-1.9) |
| Secondary | 9,035 (23.3) | 0.6 (0.4-0.9) |
| Post-secondary | 1,225 (3.2) | 0.4 (0.1-1.4) |
| **Marital status** |  |  |
| Co-habiting | 5,081 (13.1) | 1.1 (0.8-1.5) |
| Divorced/separated | 308 (0.8) | 2.1 (0.8-5.4) |
| Married | 30,446 (78.5) | 1.6 (1.3-1.9) |
| Single | 2,835 (7.3) | 0.8 (0.5-1.3) |
| Widowed | 113 (0.3) | 2.7 (1.0-7.4) |
| **Employment status** |  |  |
| Employed | 2,014 (5.2) | 0.8 (0.5-1.3) |
| Housewife | 18,956 (48.9) | 1.4 (1.2-1.7) |
| Self employed | 15,032 (38.8) | 1.6 (1.2-2.2) |
| Unemployed | 2,781 (7.2) | 1.1 (0.6-1.8) |
| **Total number of previous pregnancies** |  |  |
| 0 | 10,199 (26.3) | 0.5 (0.3-0.7) |
| 1-2 | 17,285 (44.6) | 1.2 (0.9-1.6) |
| 3-4 | 7,916 (20.4) | 2.2 (1.7-2.8) |
| >4 | 3,383 (8.7) | 3.9 (3.2-4.9) |
| **Locality** |  |  |
| Rural | 6,925 (17.9) | 2.3 (1.7-3.2) |
| Semi-urban | 13,020 (33.6) | 1.6 (1.3-2.0) |
| Urban | 18,838 (48.6) | 1.0 (0.7-1.4) |
| **Health facility level** |  |  |
| Dispensary | 6,131 (15.8) | 1.7 (1.1-2.7) |
| Health Centre | 23,837 (61.5) | 1.4 (1.1-1.8) |
| Hospital | 8,815 (8815) | 1.4 (1.0-1.9) |

### Prevalence of Syphilis among pregnant women

The overall prevalence of Syphilis infection in Tanzania among pregnant women attending ANC was 1.4% (1.2% - 1.8%). The crude prevalence of Syphilis 1.5% (0.5%- 3.8%).

The prevalence of Syphilis was the highest was among pregnant women aged 35-44 years (3.5%) and lowest (0.7%) among the youngest, 15-24 years. The burden decreased with increase in the level of education. For this, the highest burden was among pregnant women with no formal education (2.9%), while those with post-secondary education had the lowest burden (0.4%). The prevalence also increased with parity. For example, while the burden was the lowest among pregnant women in their first pregnancy (0.5%), it was the highest among those with at least five previous pregnancies (3.9%). The prevalence of syphilis was higher among women in rural areas (2.3%), compared to semi-urban (1.6%) and urban areas (1.0%). **Table 1**.

The burden varied between regions. The highest burden was in Kagera (4.5%), followed by Mbeya (3.4%), and Geita (2.7%), while the lowest prevalence was recorded in Kigoma region (0.3%). **Table 2**.

**Table 2:** Prevalence of HIV, Syphilis, and HIV& Syphilis co-infection by regions.

| Region | Total | Syphilis Infection |  |
| --- | --- | --- | --- |
|  | N | n | Prevalence (%) (95% CI) |
| Arusha | 1,542 | 11 | 0.7 (0.4-1.3) |
| Dar es Salaam | 2,874 | 19 | 0.7 (0.3-1.7) |
| Dodoma | 1,749 | 11 | 0.6 (0.2-1.8) |
| Geita | 3,460 | 93 | 2.7 (1.7-4.1) |
| Iringa | 882 | 17 | 1.9 (1.2-3.1) |
| Kagera | 1,481 | 67 | 4.5 (3-6.8) |
| Katavi | 1,224 | 20 | 1.6 (0.6-4.3) |
| Kigoma | 1,372 | 4 | 0.3 (0.1-0.8) |
| Kilimanjaro | 968 | 4 | 0.4 (0.2-0.8) |
| Lindi | 508 | 7 | 1.4 (0.7-2.9) |
| Manyara | 924 | 10 | 1.1 (0.6-2.1) |
| Mara | 1,509 | 26 | 1.7 (1.4-2.1) |
| Mbeya | 2,032 | 68 | 3.3 (2.3-4.8) |
| Morogoro | 728 | 5 | 0.7 (0.2-2.2) |
| Mtwara | 814 | 15 | 1.8 (0.9-3.6) |
| Mwanza | 2,232 | 33 | 1.5 (0.5-4.1) |
| Njombe | 670 | 17 | 2.5 (1.4-4.5) |
| Pwani | 1,841 | 8 | 0.4 (0.1-1.3) |
| Rukwa | 1,265 | 25 | 2 (1.4-2.7) |
| Ruvuma | 1,119 | 12 | 1.1 (0.3-4.1) |
| Shinyanga | 1,477 | 29 | 2 (1.3-3) |
| Simiyu | 1,956 | 24 | 1.2 (0.5-2.7) |
| Singida | 1,549 | 14 | 0.9 (0.2-3.6) |
| Songwe | 1,902 | 18 | 0.9 (0.4-2.3) |
| Tabora | 1,408 | 20 | 1.4 (0.8-2.5) |
| Tanga | 1,297 | 5 | 0.4 (0.1-1.5) |
| **Total** | **38,783** | **582** | **1.4 (1.2 - 1.8)** |

### Determinants of Syphilis infection

In a multiple logistic regression, age, education level, and the number of previous pregnancies were independent predictors of Syphilis infection among pregnant women in Tanzania mainland. Compared to younger women aged 15-24 years, women aged 25-34 years were 1.7 times more likely to test positive for syphilis infection (aOR=1.7, 95% CI:1.3-2.3), while those aged 35-44 years had about 2.8 times the odds of syphilis infection (aOR=2.8, 95%CI: 1.9-4.0) –**Table 3**.

**Table 3:** Factors associated with Syphilis infection.

| Variable | cOR (95% CI) | aOR (95% CI) | p-value |
| --- | --- | --- | --- |
| **Age group (years)** |  |  |  |
| 15-25 | Ref |  |  |
| 25-34 | 2.2 (1.7-2.9) | 1.7 (1.3-2.3) | 0.001 |
| 35-44 | 4.9 (3.6-6.6) | 2.8 (1.9-4.0) | <0.001 |
| 45+ | 3.8 (1.5-9.8) | 2.4 (1.0-5.7) | 0.042 |
| **Education level** |  |  |  |
| None | 7.8 (2.1-28.6) | 5.1 (1.3-20.2) | 0.020 |
| Primary | 4.1 (1.1-15.0) | 3.2 (0.8-12.5) | 0.100 |
| Secondary | 1.5 (0.4-4.8) | 1.4 (0.4-5.0) | 0.563 |
| Post-secondary | Ref |  |  |
| **Marital status** |  |  |  |
| Co-habiting | 0.7 (0.5-1.0) | 0.9 (0.6-1.2) | 0.410 |
| Divorced/separated | 1.3 (0.5-3.5) | 1.2 (0.4-3.2) | 0.786 |
| Married | Ref |  |  |
| Single | 0.5 (0.3-0.8) | 0.9 (0.6-1.4) | 0.696 |
| Widowed | 1.7 (0.6-5.1) | 1.2 (0.4-3.7) | 0.726 |
| **Occupation** |  |  |  |
| Employed | 0.8 (0.4-1.6) | 1.3 (0.6-2.6) | 0.474 |
| Housewife | 1.3 (0.8-2.3) | 1.2 (0.7-2.0) | 0.600 |
| Self employed | 1.5 (0.8-2.9) | 1.5 (0.8- 2.8) | 0.171 |
| Unemployed | Ref |  |  |
| **Number of previous pregnancies** |  |  |  |
| 0 | Ref |  |  |
| 1-2 | 2.5 (1.8-3.5) | 1.8 (1.2-2.5) | 0.001 |
| 3-4 | 4.7 (3.3-6.6) | 2.1 (1.4-3.1) | 0.001 |
| >4 | 8.5 (6.0-12.0) | 2.6 (1.7-3.9) | <0.001 |

| **Locality** |  |  |  |
| --- | --- | --- | --- |
| Rural | Ref |  |  |
| Semi-urban | 0.7 (0.5-1.0) | 0.8 (0.5-1.3) | 0.391 |
| Urban | 0.4 (0.2-0.7) | 0.6 (0.3-1.0) | 0.050 |
| **Health facility level** |  |  |  |
| Dispensary | Ref |  |  |
| Health Centre | 0.8 (0.5-1.4) |  |  |
| Hospital | 0.8 (0.5- 1.4) |  |  |

Compared to women with post-secondary education, women with no formal education had about five times higher odds of Syphilis infection (aOR=5.1, 95% CI: 1.3-20.2).

Further, analyses showed that, the risk of syphilis infection increased with increasing number of pregnancies. Compared with primigravids, women with 1-2 and those with 3-4 previous pregnancies had 1.8 and 2.1 higher odds of syphilis infection, (aOR=1.8, 95% CI: 1.2-2.5), and (aOR=2.1, 95% CI: 1.4-3.1). And those with more than four previous pregnancies had 2.6 times higher odds of syphilis infection compared to primigravids (aOR=2.6, 95% CI: 1.7-3.9).

Pregnant women in urban areas had 40% lower odds of Syphilis infection compared to those in rural areas (aOR=0.6, 95% CI: 0.3-1.0) –**Table 3**.

## Discussion

The results of the 2020 HIV-Syphilis sentinel surveillance among pregnant women attending ANC in Tanzania mainland reveal the prevalence of Syphilis of 1.4% (1.2% - 1.8%). This is equivalent to 30,800 (26,400- 39,600) pregnant women attending ANC in a year. There was also marked variability of the prevalence ranging from 0.3% in Kigoma to 4.5% in Kagera regions. Our results also indicate that higher Syphilis infection among pregnant women are associated with older age, no education, and increasing number of pregnancies.

Syphilis being prevalent among 1.4% pregnant women in Tanzania is worrisome, owing to the unprecedented impact the disease can have to the women and birth outcomes. The national prevalence of syphilis in this survey is in-line with the decreasing trend of Syphilis among pregnant women as the national prevalence of Syphilis for pregnant women has continued to decline from 4.3% in 2008, to 2.5% in 2011, and to 1.8% in 2017 (Swai *et al*., 2006; Manyahi *et al*., 2015). Further, compared to the estimated prevalence of Syphilis among pregnant women in the East African region, 3.2%, and that of the African region 2.5%, the prevalence is lower in Tanzania (Hussen and Tadesse, 2019). Tanzania has maintained the downward trend of the prevalence for pregnant women over the years. However, the prevalence is still much higher than the global average of Syphilis among pregnant women, 0.6% (Korenromp *et al*., 2019). To successfully eliminate Syphilis concerted efforts are still needed to address syphilis infection among pregnant women.

In this study, the prevalence of syphilis was higher among older pregnant women than younger pregnant women and also lowest among primegravids women and increased with the number of pregnancies. Untreated Syphilis can evolve from primary stage and secondary stages to latent stage where the symptomatology of the secondary stage resolves. As these women will still have a positive serologic test but lack clinical manifestation, it is possible that the increased prevalence in older women than younger ones and among multipara women than primegravids indicate that many pregnant women in Tanzania remain untreated and have a latent Syphilis or are re-infected as their partners remain untreated (Tsimis and Sheffield, 2017). And although at this stage patients are not considered infectious through sexual transmission, vertical transmissions have been reported (Tsimis and Sheffield, 2017). Treatment for Syphilis is available in the country, therefore efforts to screen pregnant women and their partners should be emphasized and linking both to care if positive. The consequences of untreated syphilis to the unborn baby include stillbirth, and congenital syphilis among others (Gomez *et al*., 2013; Shava, Moyo and Zash, 2019). Syphilis also facilitate the vertical transmission of HIV, an existent concern in settings with high HIV burden such as Tanzania (Mwapasa *et al*., 2006).

In this study, education level was associated with Syphilis infection among pregnant women where women who had no education had significantly higher prevalence of Syphilis than women with higher education levels. While education was significantly associated with syphilis in the 2003/2004 surveillance of pregnant women in Tanzania, it was not in the 2011 surveillance (Swai *et al*., 2006; Manyahi *et al*., 2015). This may be due to the scaling up of the Syphilis screening and testing since the WHO declaration to eliminate Syphilis in 2008 (WHO, 2021). The association of Syphilis and education has also been reported among women and men in the general population in Kenya (Otieno-Nyunya *et al*., 2011). Having education renders one less likely to engage in risky sexual behaviors, but enables access and uptake to care and treatment (Leon *et al*., 2017). There was also borderline evidence of higher Syphilis infection in rural areas compared to urban areas.

The interpretation of these results is not without a potential limitation. Although the survey spanned across all the 26 regions of Tanzania mainland, the generalizability of the findings should be with caution. Most of the sites used in this survey were inherited from previous surveys and there have been many newer PMTCT sites which could have featured in the sampling frame had the sampling conducted again. Thus, the choice to inherit previous PMTCT sites might have narrowed our potency to be as representative as possible. Moreover, this is a cross sectional study whose findings may not ascertain causality. However, the survey offer a comparison with previous surveys, and trends can be established. With such high sample size, the study provides a more reliable estimate of national prevalence, with sub-national estimates adequate for interventional planning and execution.

## Conclusion

Syphilis remains prevalent among pregnant women in Tanzania with wide regional burden disparity. Efforts to prevent new infections, screen pregnant women, and treat those infected should be strategized to include all regions and renewed emphasis in regions with high burden, and importantly among women who are multipara, with the low level of education, and advanced age.

## Data Availability

We cannot at this time share the de-identified data file with the third party owing to the Tanzania Health Research Regulation that requires the third party to sign the Data Transfer Agreement. Data may be shared upon request by any researcher to the Ministry of Health (the owner) and upon fulfilling the regulatory requirements.

## Acknowledgment

We wish to thank all regional and district medical officers, regional and district reproductive, and sexual health coordinators for their support in the implementation of the survey. Our sincere appreciations to the nurses at the facilities for their commitment throughout the study.

